# Neovascular Glaucoma at a Tertiary Centre in Finland, 2008–2024: A Retrospective Cohort Study

**DOI:** 10.64898/2026.06.01.26354330

**Authors:** Gilbert Simons, Mikael von Fersen, Paula Summanen, Mika Harju

## Abstract

**Background/Aims:** Neovascular glaucoma (NVG) is an aggressive secondary glaucoma with limited longitudinal data. This study reports the aetiologies, treatments, and longitudinal outcomes in NVG.

**Methods:** Patients with NVG were identified through electronic medical record review. Inclusion required documented rubeosis of the iris and/or anterior chamber angle, intraocular pressure (IOP) ≥25 mmHg, diagnosis during 2008–2024, and follow-up at Helsinki University Hospital. Baseline data and all follow-up visits were included.

**Results:** Of 919 patients identified, 626 met inclusion criteria, with a median follow-up of 24 months. The estimated NVG incidence was 2.2/100,000/year. The most common aetiology was central retinal vein occlusion (CRVO; 45%), followed by diabetic retinopathy (DR; 14%), central retinal artery occlusion (CRAO; 11%), and ocular ischaemic syndrome (8%).

Half of patients had hand motion vision or worse at baseline, with 18% at no light perception (NLP). At 5 years, 13% had Snellen 6/60 vision or better. Visual outcomes differed by aetiology, with median time to NLP ranging from 1.6 (CRAO) to 9.1 (DR) years (log-rank *p*=0.002). Median baseline IOP was 40 mmHg, decreasing to 21 mmHg by 1 year. Ocular pain fell from 43% at baseline to 11% at last follow-up. Structural eye loss (e.g., enucleation or phthisis) occurred in 3% by 5 years.

**Conclusion:** The estimated NVG incidence was lower than previously reported elsewhere. Unlike other cohorts where DR predominates, CRVO was the most common aetiology, and visual prognosis was strongly aetiology-dependent. Glaucoma drainage device surgery reached 7.6% at 3 years, despite the severity and refractory nature of NVG.

## Introduction

Neovascular glaucoma (NVG) is a rare form of secondary glaucoma that can lead to severe and irreversible vision loss. In a nationwide cohort study from Taiwan, the age-standardised incidence rate of NVG was reported as 4.33 cases per 100,000 person-years [1]. A 2005 retrospective analysis described 3.9% of all glaucoma cases in Craiova, Romania, as the neovascular type [2]. Aside from these reports, the incidence and prevalence of NVG have rarely been described, and never in Finland.

The four conditions most commonly predisposing to NVG are all of ischaemic origin: diabetic retinopathy (DR), ischaemic central retinal vein obstruction (CRVO), ocular ischaemic syndrome (OIS) and central retinal artery occlusion (CRAO) [3-5]. During ischaemia, vascular endothelial growth factor (VEGF) is secreted, amongst other factors, initiating the growth of fragile, new vessels. These abnormal vessels are often first noticed at the pupillary margin of the iris, causing rubeosis iridis [6]. Subsequently, the process spreads to the trabecular meshwork where angiogenesis occurs among contracting myofibroblasts, hindering outflow of aqueous humour, often elevating intraocular pressure (IOP) [3, 7]. Other clinical manifestations include peripheral anterior synechiae formation resulting in secondary angle-closure glaucoma [7].

NVG is often refractory to IOP-lowering medication. Without treating the underlying cause and therefore reducing angiogenic factors such as VEGF in the aqueous humour, the neovascularisation progresses [3, 8]. The European Glaucoma Society divides the management of NVG into two approaches. The first targets the underlying cause with anti-VEGF injections and pan-retinal photocoagulation (PRP). The second addresses the glaucoma itself with topical and systemic medications, as well as surgical interventions [9].

Retrospective NVG studies have been conducted, including large cohorts, but rarely in European populations and often in selected ones. Many are also limited to first and last visits, rather than the clinical course between diagnosis and outcome, and few analyse aetiologies separately [1, 10-13]. This study was designed to address these gaps by reporting the aetiologies, treatments, and longitudinal clinical outcomes across all visits of all NVG patients treated at the Department of Ophthalmology, Helsinki University Hospital, between 2008 and 2024.

## Materials and methods

### Study design

This was a retrospective, descriptive cohort study including all NVG patients treated at HUS Helsinki University Hospital, Finland, between 2008 and 2024. Reporting follows the STROBE statement [14]. Patients with the ICD-10 diagnosis H40.5 and a mention of NVG (or its Finnish-language inflected forms) in any ophthalmologic electronic medical records (EMR) were identified, and all their ophthalmologic records were retrieved [15]. Complete diagnosis and medication lists were retrieved, as was date of death where applicable. Two trained reviewers independently analysed non-overlapping subsets of the EMRs during 2025–2026.

### Inclusion criteria

Inclusion required documented rubeosis of the iris and/or anterior chamber angle, together with IOP ≥25 mmHg recorded at least once in the EMR. The IOP threshold ensured inclusion of NVG cases requiring treatment.

The date of diagnosis was defined as the first visit meeting inclusion criteria. Additional requirements included diagnosis made during 2008–2024, treatments and follow-ups primarily at HUS, and at least two records on separate dates. Only one eye per patient was included: the first diagnosed, or the eye with higher IOP if both were diagnosed simultaneously. All records from diagnosis to 31 December 2024 were analysed. Follow-up reflected routine clinical care without a protocol-defined visit schedule. All ophthalmologic records of the affected eye were included, irrespective of subspecialty. Sample size was not pre-specified; all eligible patients were included.

HUS served roughly 32% of the Finnish population at the end of 2024 [16], and was the only tertiary centre in the area. The centre included one central ophthalmologic department and two smaller units. Some patients were also referred to outsourced services for follow-up. These records were incrementally available from July 2022 onward.

### Baseline data

Baseline data extracted from EMRs, diagnosis lists, and medication records included prior ocular and systemic diseases, glaucoma subtypes, medications, ocular surgery, and NVG aetiology. Prior systemic diseases included diabetes (with its type), hypertension, dyslipidaemia and peripheral artery disease. The most prevalent glaucoma subtypes in the cohort were retained as separate categories: primary open-angle, exfoliation, and normal-tension glaucoma; remaining subtypes were pooled as ‘other’. Systemic medications collected were insulin and antithrombotic agents. To accommodate delayed EMR documentation, pre-existing conditions recorded up to six months after diagnosis, and medications up to three months after, were included as baseline.

Aetiology was assigned on EMR review. CRVO and CRAO required fundus-visible occlusion. DR required retinopathy judged clinically responsible for the neovascular process. OIS required consistent fundus findings together with significant carotid artery stenosis on ultrasound. When multiple causes were identified, the dominant was recorded as primary and the additional as secondary. Uncommon aetiologies were pooled and cases without an identifiable cause recorded as undefined.

### Visit data

At each visit, visual acuity (VA), IOP, pain status, biomicroscopic findings, and administered treatments were recorded. Best-corrected VA (BCVA) was recorded and categorised as numerical values, Early Treatment Diabetic Retinopathy Study (ETDRS) letters, counting fingers (CF), hand motion (HM), light perception (LP), or no light perception (NLP). Partly read decimal acuity lines were rounded down to the next full line and plus notations were ignored. Chart acuities were converted to the logarithm of the minimum angle of resolution (logMAR = −log_10_(decimal acuity)); ETDRS as logMAR = (85 − letters)/50. LP and NLP were fixed at 2.7 and 3.0 [17]; CF and HM were mapped to 2.05–2.30 and 2.35–2.60. When distance was recorded, interpolation within the interval was applied; otherwise, the midpoint was used. A distance of ‘ad oculum’ was treated as 0.10 m. For IOP, Goldmann applanation tonometry (GAT) was preferred when available, with the iCare rebound tonometer (Icare Finland Oy, Vantaa, Finland) as fallback. Rare measurements by other methods were pooled with rebound tonometry. IOP was recorded before chemical mydriasis when possible. Pain status was categorised as absent, mild, or severe. Only ocular pain or aching was included; other non-specific sensations were excluded.

Recorded treatments included: topical and systemic glaucoma medications by active substance (prostaglandins, beta blockers, topical and systemic carbonic anhydrase inhibitors, alpha agonists, parasympathomimetics, and Rho-kinase inhibitors), Scatter retinal photocoagulation (number of spots), anti-VEGF intravitreal injections, transscleral cyclophotocoagulation (TSCPC), peripheral retinal cryotherapy, glaucoma drainage devices (GDDs), and other surgical procedures. The biomicroscope, visual field and optical coherence tomography data are described in Supplementary Methods.

### Primary outcomes

Primary outcomes were BCVA, pain status, and IOP. The four main aetiologies, CRVO, DR, CRAO, and OIS, were compared for differences in baseline characteristics and clinical outcomes. Fixed timepoints of baseline, 1 year, 3 years, and 5 years were selected, with windows of ±30, ±90, ±180, and ±180 days, respectively. The baseline timepoint was constructed from the visit at which the diagnostic criteria were met. If individual measurements were absent, a second visit within 30 days was used to supplement missing values. For all other timepoints, the single closest visit within the respective window was selected. For pain status, the last recorded visit was used as the final timepoint since documentation at later fixed intervals was too sparse for aetiology-stratified reporting.

### Statistical analysis

Statistical analysis was performed using Python (version 3.12.10, Python Software Foundation, Wilmington, DE, USA) [18, 19]. ChatGPT and Claude (OpenAI and Anthropic, San Francisco, CA, USA) [20, 21] assisted in drafting and debugging the analysis code, with all output verified against the source data by the lead author. Continuous variables were compared across aetiologies using the Kruskal–Wallis test, with Mann–Whitney U post-hoc tests where significant. Categorical variables were compared using the chi-square test, with Fisher’s exact (2×2) or chi-square (larger tables) post-hoc tests where significant. Fixed-timepoint comparisons required at least five observations per group. All between-group testing was exploratory, with no correction for multiple comparisons. Time-to-event outcomes used Kaplan–Meier estimation, with aetiology differences assessed by log-rank test.

Patients without a recorded visit within a given timepoint window were excluded from that timepoint’s analysis. In time-to-event analyses, censoring was at the last visit or death, whichever came first. A *p*-value <0.05 was considered statistically significant. Per the Finnish Social and Health Data Permit Authority (Findata) regulations, groups with fewer than three observations were suppressed or pooled.

### Study approval

The study was approved by HUS Institutional Review Board (HUS/20/2025) and followed the guidelines of the Declaration of Helsinki. Registry studies are not classified as medical research by current Finnish legislation, and the study therefore did not require a separate ethics approval.

## Results

### Included patients and neovascular glaucoma incidence

A total of 919 patients were identified, of whom 626 met the inclusion criteria. The most common reasons for exclusion included diagnosis prior to 2008, follow-up at other hospital districts, absence of rubeosis, or IOP below threshold (Figure 1).

**Figure 1.**
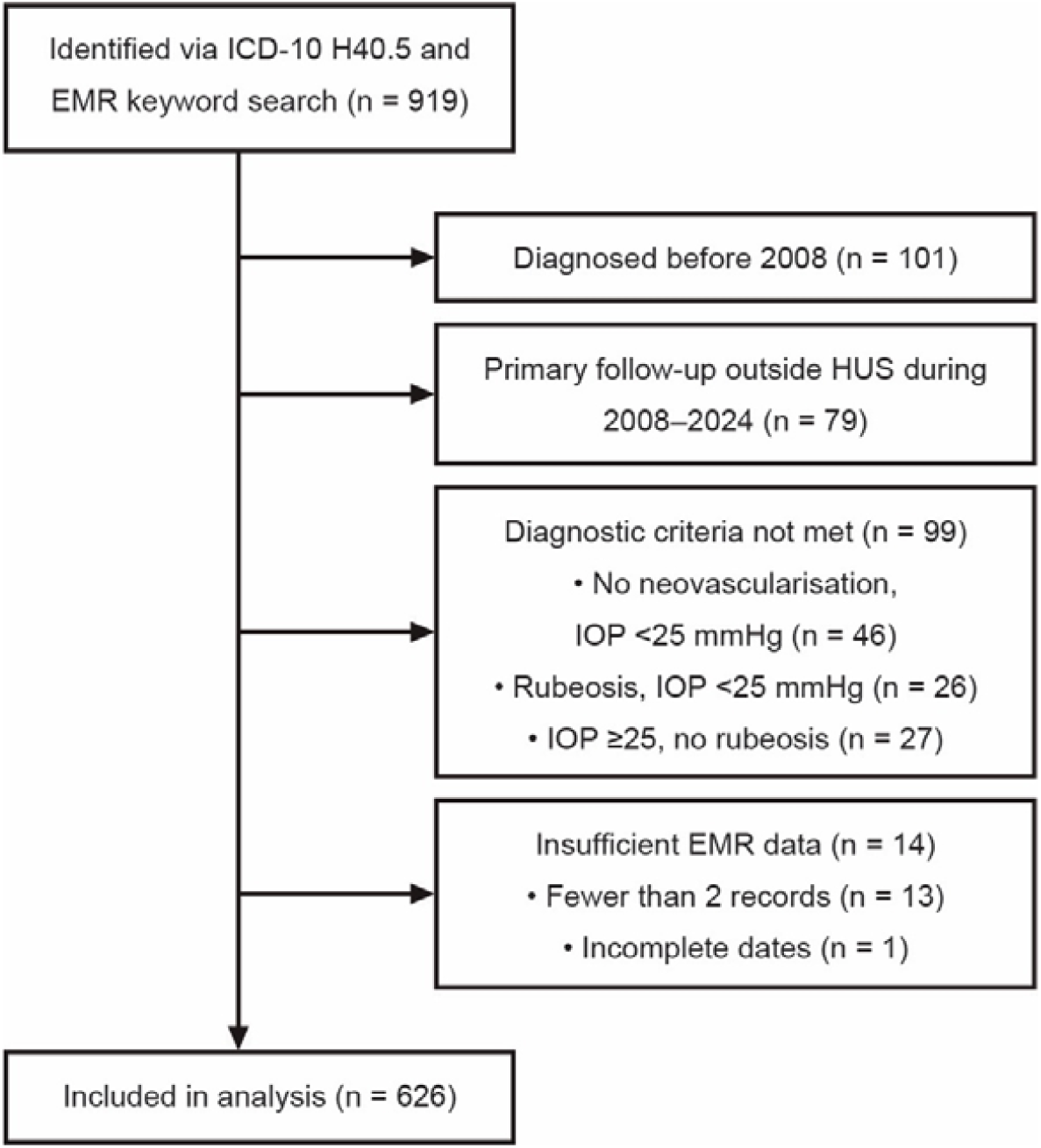
Inclusion criteria. 919 patients were identified, of whom 626 were included in the study. The most common reasons for exclusion were diagnosis before 2008, primary follow-up outside HUS Helsinki University Hospital, and not meeting diagnostic criteria. EMR, electronic medical record; IOP, intraocular pressure.

A mean of 36.8 (standard deviation 7.3) new NVG cases were diagnosed annually at HUS, corresponding to an estimated incidence of 2.2 per 100,000 in the catchment area using the 2016 HUS-area population as the mid-period denominator [16]. NVG accounted for approximately 2.4% of new glaucoma cases annually, based on new medication reimbursement entitlements in the HUS area [22]. There was no clear trend in annual incidence across the study period (Supplementary Figure 1).

### Patient demographics and aetiologies

Patients had a median age of 76 (interquartile range, IQR 66–84) years at diagnosis. Sex was divided as 352 (56%) male and 274 (44%) female patients. Median visits per patient was 14 (9–22), and median follow-up 24 (9–58) months. The most common aetiology was CRVO with 283 patients (45%), followed by DR in 90 cases (14%), CRAO in 66 (11%) and OIS in 49 (8%).

Comparing aetiologies revealed that CRVO had the largest proportion of prior glaucoma (*p*<0.001). Additionally, DR patients were younger, had longer follow-up, and accumulated more visits than the other aetiology groups (*p*<0.001 for all three). Table 1 and Supplementary Tables 1 and 2 provide further demographics, systemic history, and baseline biomicroscopic findings by aetiology, with post-hoc results in Supplementary Table 3.

**Table 1.** Patient demographics, aetiologies, systemic and ocular history.

| **Cohort characteristics —**<br>**continuous variables: median [IQR]; categorical: n (%)** | **Baseline** |
| --- | --- |
| Total patients, n | 626 |
| Age at diagnosis, years | 76 [66–84] |
| Follow-up, months | 24 [9–58] |
| Visits per patient | 14 [9–22] |
| **Sex** |  |
| Male | 352 (56%) |
| Female | 274 (44%) |
| **Affected eye** |  |
| OD | 317 (51%) |
| OS | 309 (49%) |
| Bilateral neovascular glaucoma | 28 (4%) |
| **Aetiology** |  |
| Central retinal vein occlusion | 283 (45%) |
| Diabetic retinopathy | 90 (14%) |
| Central retinal artery occlusion | 66 (11%) |
| Ocular ischaemic syndrome | 49 (8%) |
| Retinal detachment | 18 (3%) |
| Uveal melanoma | 13 (2%) |
| Branch retinal vein occlusion | 9 (1%) |
| Branch retinal artery occlusion | 3 (0.5%) |
| Ocular lymphoma | 3 (0.5%) |
| Undefined aetiology | 76 (12%) |
| Other (individual n<3) | 16 (2.6%) |
| Secondary aetiology registered | 27 (4.3%) |
| **Systemic comorbidities** |  |
| Diabetes mellitus | 266 (42%) |
| Type 1 | 42 (7%) |
| Type 2 | 215 (34%) |
| Other/undefined type | 9 (1%) |
| Hypertension | 382 (61%) |
| Dyslipidaemia | 198 (32%) |
| Peripheral artery disease | 82 (13%) |
| **Prior glaucoma** |  |
| Glaucoma | 174 (28%) |
| Primary open-angle glaucoma | 79 (13%) |
| Exfoliation glaucoma | 78 (12%) |
| Normal-tension glaucoma | 4 (1%) |
| Other glaucoma subtype | 13 (2%) |
| **Prior ocular treatment<sup>1</sup>** |  |
| Anti-VEGF injections | 71 (11%) |
| Scatter retinal photocoagulation | 94 (15%) |
| Vitrectomy | 62 (10%) |
| Laser iridotomy | 27 (4%) |
| Selective laser trabeculoplasty | 17 (3%) |
| Glaucoma filtration surgery | 15 (2%) |
| Peripheral retinal cryotherapy | 5 (1%) |

<sup>1</sup>Any procedure or injection recorded before the date of NVG diagnosis. Prior ocular treatments are reported as presence or absence only; quantitative details, such as scatter retinal photocoagulation spot counts, were not
captured. Scatter retinal photocoagulation includes panretinal photocoagulation and less extensive retinal laser treatments. IQR, interquartile range; VEGF, vascular endothelial growth factor.

### Visual acuity

Baseline median BCVA was 2.5 (IQR 2.1–2.7) logMAR, with 110 (18%) patients at NLP. Median BCVA was 2.7 (1.2–3.0) logMAR at 5 years. When analysing aetiologies separately, outcomes diverged significantly at all time points (*p*<0.001– 0.004, pairwise comparisons in Supplementary Table 3), with DR achieving the best outcomes. The difference was sustained across follow-up as shown by survival analysis (log-rank *p*=0.002; see Kaplan–Meier section). The proportion of patients declining to NLP within three years of diagnosis showed a modest increasing trend over the study period from approximately 40% in the earliest years to around 50– 60% towards the end. See Figure 2 for BCVA per aetiology at different timepoints as well as the proportion of patients declining to NLP.

**Figure 2.**
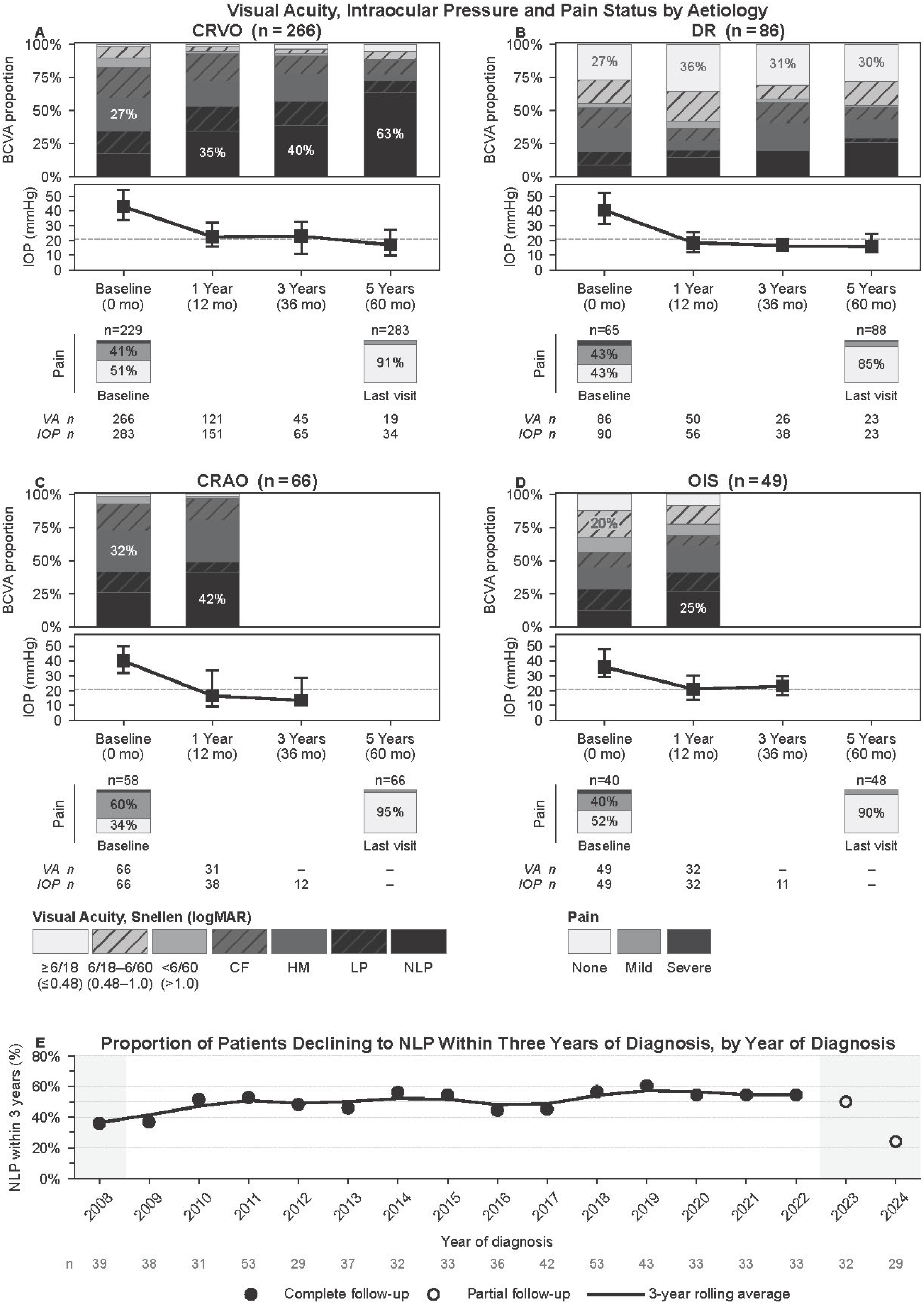
Visual acuity, intraocular pressure, and pain status by aetiology over time. Panels A–D show BCVA, IOP, and pain outcomes for the four main aetiologies (A: CRVO, B: DR, C: CRAO, D: OIS). Stacked bars represent the proportion of eyes in each BCVA category at baseline, 1 year, 3 years, and 5 years, ordered from best (light, top) to worst (black, bottom), with the largest segment labelled. Chart BCVA was divided into three categories following WHO ICD-11 classification [23]: ≥6/18, mild impairment or better (logMAR ≤0.48), 6/18 to 6/60, moderate visual impairment (logMAR 0.48–1.0), and worse than 6/60, severe visual impairment or worse (logMAR >1.0) to the chart limit. Off-chart acuities were reported separately as CF, HM, LP, and NLP. The stacked bars have a slight jitter, to protect anonymity amongst groups with small counts. The line plot shows median IOP with IQR error bars; the dashed line indicates 21 mmHg. The lowest element per panel shows the proportion of eyes in each pain category at baseline and at the last recorded visit. The n-at-risk rows show eyes with available BCVA and IOP measurements at each timepoint. Aetiology counts differ slightly from Table 1 due to missing baseline BCVA values (n=35). Results of groups of 10 or fewer are omitted to preserve anonymity. Panel E shows the proportion of patients declining to NLP within 3 years of diagnosis, plotted by year of diagnosis. Dots represent annual data and lines a rolling 3-year average. Hollow dots indicate patients diagnosed in 2023–2024 without complete 3-year follow-up; 2008 is incomplete for the rolling average. BCVA, best-corrected visual acuity; IOP, intraocular pressure; IQR, interquartile range; logMAR, logarithm of the minimum angle of resolution; CF, counting fingers; HM, hand motion; LP, light perception; NLP, no light perception; WHO, World Health Organization.

### Intraocular pressure and pain

Across all available baseline measurements, median IOP was 40.0 (IQR 32.0–51.0) mmHg by GAT and 40.0 (30.0–50.0) mmHg by rebound tonometry. GAT was recorded in 546 patients (87%) and rebound tonometry in 435 (70%), with both methods in 355 (57%). IOP decreased for all aetiologies during the first year, stabilising near 21 mmHg. Ocular pain was present in 270 patients (43%) at baseline, decreasing to 11% at the last recorded value. The difference between groups for GAT and pain was not statistically significant. IOP and pain at different timepoints per aetiology can be seen in Figure 2.

### Treatments after diagnosis

PRP (cumulative threshold ≥4000 spots) was performed in 35% of patients at 1 year and 37% at 3 years. Applying a lower threshold of 250 spots gave a median of 15 (95% confidence interval, CI 10–22) days to treatment. Peripheral retinal cryotherapy was given to 51% at 1 year and 60% at 3 years (median 299 days; 95% CI 170–730 days).

Anti-VEGF injections were given to 32% at 1 year and 36% at 3 years, totalling 742 injections in 200 patients. TSCPC was performed in 56% of patients by 1 year and 64% by 3 years (median 177 days; 95% CI 122–298 days). Surgery with GDD implantation was done in 4.4% of patients at 1 year and 7.6% at 3 years; 32 unique patients received one.

At baseline, 7.3% of patients used no IOP-lowering medication and 42% used more than three agents, compared with 18% and 29% at 3 years. Over the study period, anti-VEGF use increased and the proportion of patients accumulating ≥4000 PRP spots within 3 years declined. Cumulative treatment uptakes between aetiologies and their trends over time are illustrated in Figure 3. Supplementary Figure 2 shows cumulative PRP spots per patient by diagnosis year.

**Figure 3.**
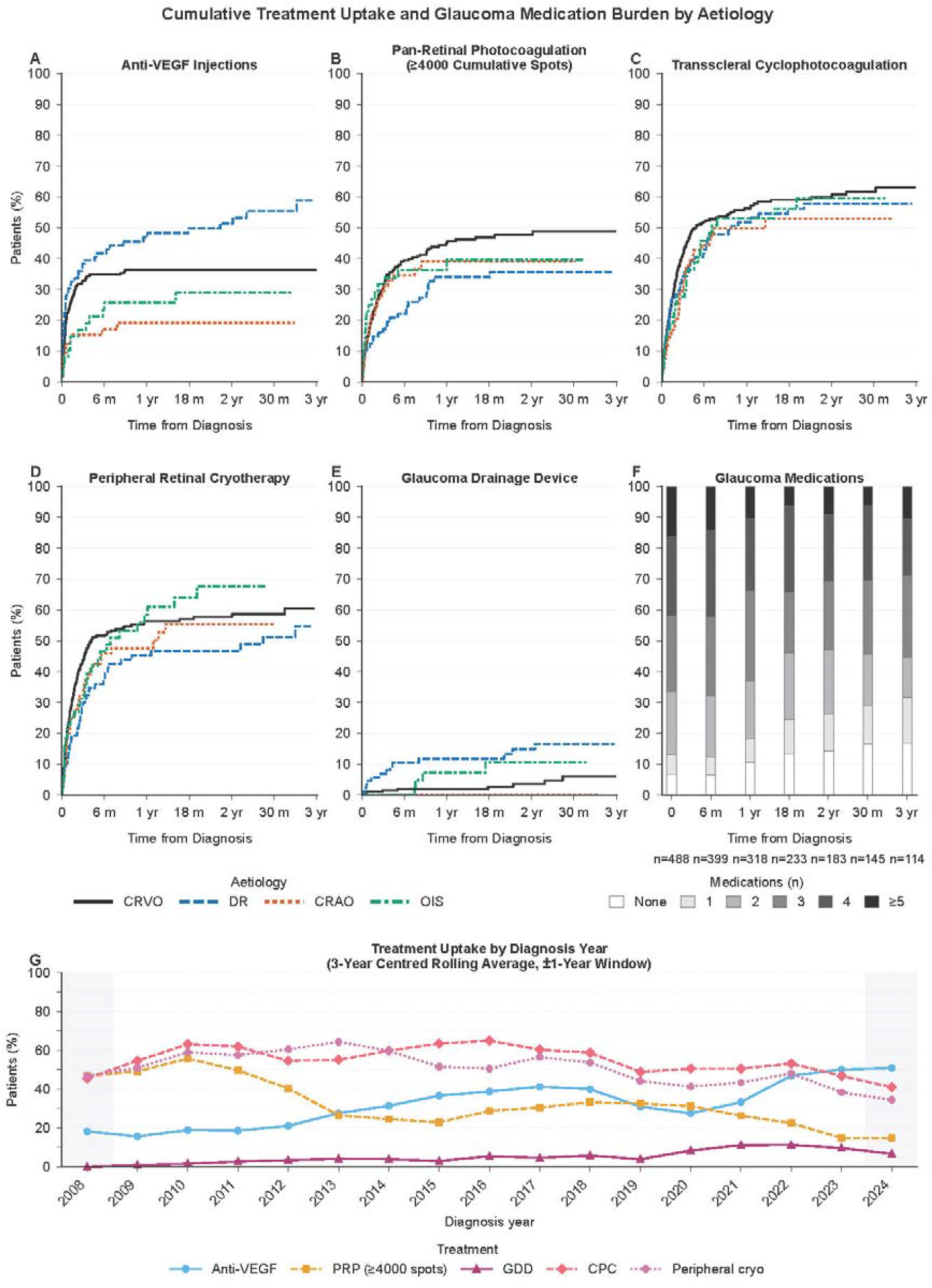
Cumulative treatment uptake and glaucoma medication burden by aetiology. Panels A–E show Kaplan–Meier cumulative incidence curves for time to first treatment from diagnosis. Each patient contributes one event, defined as the first visit at which the treatment was recorded. Baseline treatments were excluded. A cumulative minimum of 4000 spots were required to qualify for pan-retinal photocoagulation. Patients without the treatment within the observation window were censored at their last visit. Curves are truncated where fewer than ten patients remained at risk to preserve anonymity. The x-axes were limited to 3 years since truncation started appearing and curves flattened out. Panel F shows the distribution of the number of topical and systemic glaucoma medications in use at discrete timepoints, displayed as proportions CRVO, DR, CRAO and OIS patients with a visit within the timepoint window. Glaucoma medications were prostaglandins, beta blockers, topical and systemic carbonic anhydrase inhibitors, alpha agonists, parasympathomimetics, and Rho-kinase inhibitors. The number of contributing patients is shown below each bar. Panel G shows the three-year average of treatment trends as a function of time. Anti-VEGF, anti-vascular endothelial growth factor; CRVO, central retinal vein occlusion; DR, diabetic retinopathy; CRAO, central retinal artery occlusion; OIS, ocular ischaemic syndrome.

### Kaplan–Meier survival analysis

Kaplan–Meier analysis (Figure 4) showed that the probability of maintaining vision above NLP was 70% (95% CI 65–74%) at 1 year, 56% (50–61%) at 3 years, and 47% (41–54%) at 5 years. Deterioration to NLP differed statistically significantly across aetiologies (log-rank *p*=0.002), with CRVO and CRAO showing the steepest declines. DR and OIS retained light perception longer, with median times to NLP of 9.1 (5.7–not reached) and 6.3 (3.7–not reached) years, compared with 3.5 (1.9–6.4) years for CRVO and 1.6 (0.6–3.7) years for CRAO. The overall median was 4.6 (2.9–7.8) years.

**Figure 4.**
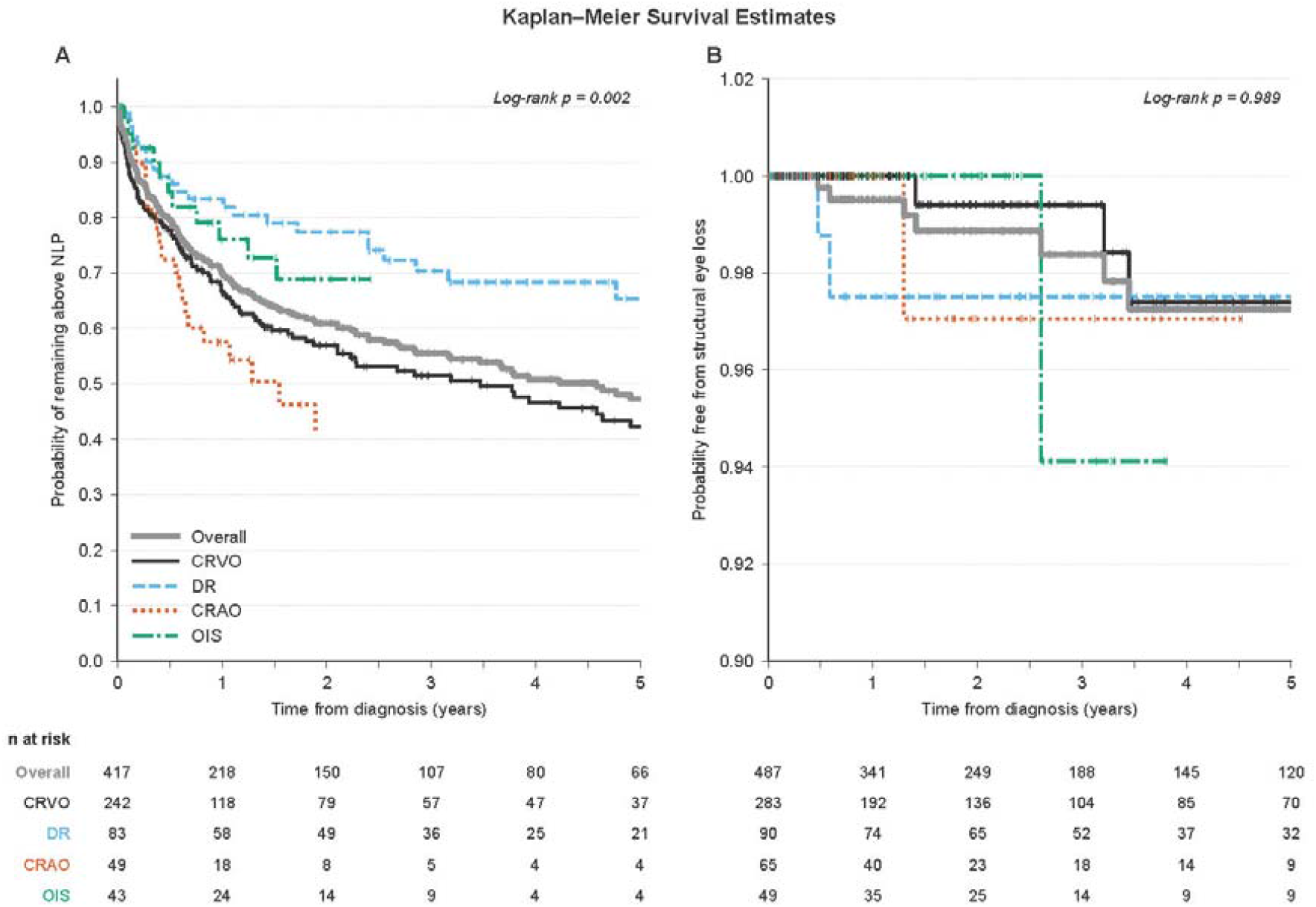
Kaplan–Meier survival estimates by aetiology. Panel A shows the probability of remaining above no light perception (NLP) from the time of diagnosis, excluding patients already at NLP at baseline. Panel B shows the probability of remaining free from terminal eye loss, defined as evisceration, enucleation, alcohol injection, phthisis developing after diagnosis, or shell prosthesis. Curves are shown for the four main aetiologies and the overall cohort. Tick marks indicate censored observations. Curves are truncated when fewer than ten patients remain at risk to protect anonymity. Differences between aetiology groups were assessed using the multivariate log-rank test. CRVO, central retinal vein occlusion; DR, diabetic retinopathy; CRAO, central retinal artery occlusion; OIS, ocular ischaemic syndrome.

Structural eye loss, comprising enucleation, evisceration, alcohol injection, new-onset phthisis, and shell prosthesis fitting, had occurred in 2.7% of patients at 5 years (95% CI 1.3–5.9%). There was no significant difference between aetiologies (log-rank *p*=0.989; Figure 4).

## Discussion

This was a retrospective cohort study of all NVG patients treated at HUS between 2008 and 2024. The annual incidence was lower than previously reported [1, 2] and showed no clear upward trend over the study period. The aetiology distribution differed from many previous studies, with CRVO rather than DR as the dominant cause [1, 2, 11, 24, 25]. These two findings may be related: the rise of diabetes has been proposed as a driver of increasing NVG incidence, yet Finland’s national free-of-charge DR screening programme has reduced the rate of sight-threatening complications from DR, likely limiting its contribution to NVG [26].

The DR group had significantly better VA outcomes than the other aetiologies, consistent with Nakano *et al*. [27]. Survival analysis showed that deterioration to NLP was steep overall, with CRVO and CRAO declining most rapidly and DR showing the slowest decline. To our knowledge, no comparable study has reported aetiology-stratified Kaplan–Meier curves for time to NLP in an unselected NVG cohort, making direct numerical comparison difficult. The overall probability of structural eye loss was low throughout follow-up, suggesting that cosmetically acceptable outcomes are achievable even when visual prognosis is poor.

Median IOP decreased from 40 mmHg at baseline to 21 mmHg at 1 year, and 17 mmHg at 5 years, consistent with the reduction reported by Liao *et al*. [10], although their cohort relied predominantly on GDDs rather than the mixed approach used in the present study. Their patients required fewer glaucoma medications after intervention than ours did. Nakano *et al*. [27] aimed to treat their patients with PRP and trabeculectomy, resulting in higher final IOP in RVO and OIS patients than in our cohort, whereas DR outcomes were comparable. Moreover, pain has been sparsely reported longitudinally. Strzalkowski *et al*. [28] reported similar baseline pain with pain-free follow-ups under a standardised treatment protocol. We report overall 11% pain at last recorded pain value. For DR, this was higher at 15%.

Published treatment data are difficult to compare, as many previous studies draw on selected populations and are weighted towards surgical approaches [24, 29-32]. Kingston *et al*. [25] reported a considerably higher proportion receiving GDD treatments than the present study, whereas TSCPC was similar, although their cohort was substantially smaller. Post-diagnosis PRP uptake is underestimated, as pre-diagnosis scatter laser photocoagulation was recorded only as present or absent. Top-up sessions below the 4000-spot threshold went uncaptured, particularly in DR patients (67% pre-treated). The declining use of PRP alongside increased anti-VEGF uptake over the study period likely reflects the progressive adoption of anti-VEGF agents into NVG management guidelines. PRP is in any case often unfeasible in this predominantly elderly cohort with frequent media opacity.

CRAO as a distinct NVG subgroup has rarely been reported in the literature. In the present cohort, CRAO had the highest proportion of patients at NLP at baseline. Despite an initially slower VA decline than CRVO, CRAO showed a steeper trajectory beyond six months, ultimately reaching similarly poor visual outcomes.

A key strength of this study is the minimal selection bias. All patients were included regardless of presenting VA, inpatient or outpatient status, treatment received, or follow-up duration (requiring at least one follow-up visit). As the sole tertiary ophthalmological centre in the region, HUS covers a large and geographically defined catchment area. The patient identification process was deliberately broad, capturing suspected as well as confirmed NVG cases and reducing the risk of underascertainment.

Another strength of the study was the cohort size. It allowed for aetiology-stratified analysis, which to our knowledge has not been performed on this scale in a European population. Inclusion of all visits rather than only first and last enabled longitudinal characterisation of outcomes across the full follow-up period. Limitations include the retrospective design and the inherent documentation bias of EMR-based data collection, affecting both the treating clinicians and the researchers extracting data. Aetiology was determined by clinical judgement, with uncertain cases recorded separately. The IOP inclusion threshold of 25 mmHg excluded some milder cases, though these numbered only 22 patients. Statistical disclosure requirements under the Findata permit precluded reporting of subgroup cells with fewer than three observations, restricting some aetiology-stratified analyses at later timepoints. Follow-up data were also sparse at later intervals, partly because EMRs from outsourced follow-up services were only available from mid-2022 onward. The cohort is predominantly Finnish-speaking and elderly, reflecting the Finnish population, which may limit generalisability to other ethnic groups [33].

## Conclusion

This is among the few NVG cohorts reported from Europe, and one of few anywhere with longitudinal data. CRVO was the dominant aetiology, in contrast to the DR predominance seen in many previous studies, and DR patients achieved the best visual outcomes. The incidence of NVG was lower than previously reported, reflecting fewer DR patients. To our knowledge, this is among the first studies to analyse CRAO separately. These findings provide a foundation for improving the management and prognosis of this severe disease.

## Declaration Statements

## Acknowledgements

Preparing this work, the lead author (GS) used ChatGPT and Claude (OpenAI and Anthropic, San Francisco, CA, USA) [20, 21]. These tools were used to improve the cohesion and coherence of the language. Their use in drafting and debugging the analysis code is described in the Methods. The tools were not used to design the study, select statistical methods, analyse data, or interpret findings. The authors take full responsibility for the content.

## Contributors

GS: Conceptualisation, Data curation, Funding Acquisition, Formal analysis, Investigation, Methodology, Software, Validation, Visualisation, Writing – original draft. MvF: Data curation. PS: Conceptualisation, Supervision, Writing – review & editing. MH: Conceptualisation, Funding Acquisition, Methodology, Supervision, Writing – review & editing.

## Ethics approval

The study was approved by HUS Helsinki University Hospital Institutional Review Board (HUS/20/2025). In accordance with the Finnish Act on the Secondary Use of Health and Social Data (552/2019) and the research permit, individual patient consent or separate ethical review was not required. The data was pseudonymised.

## Data availability statement

All data produced during this study are included in this published article or supplementary information. Due to data protection regulation, patient-level data cannot be shared.

## References

1. Lin PA, Lee CY, Huang FC, Huang JY, Hung JH, Yang SF. Trend of Neovascular Glaucoma in Taiwan: A 15-year Nationwide Population-based Cohort Study. Ophthalmic Epidemiol. 2020;27(5):390–8.

2. Mocanu C, Barăscu D, Marinescu F, Lăcrăţeanu M, Iliuşi F, Simionescu C. [Neovascular glaucoma--retrospective study]. Oftalmologia. 2005;49(4):58–65.

3. Rodrigues GB, Abe RY, Zangalli C, Sodre SL, Donini FA, Costa DC, et al. Neovascular glaucoma: a review. International Journal of Retina and Vitreous. 2016;2(1):26.

4. Brown GC, Magargal LE, Schachat A, Shah H. Neovascular glaucoma. Etiologic considerations. Ophthalmology. 1984;91(4):315–20.

5. Yang H, Yu X, Sun X. Neovascular glaucoma: Handling in the future. Taiwan J Ophthalmol. 2018;8(2):60–6.

6. Laatikainen L, Blach RK. Behaviour of the iris vasculature in central retinal vein occlusion: a fluorescein angiographic study of the vascular response of the retina and the iris. Br J Ophthalmol. 1977;61(4):272–7.

7. John T, Sassani JW, Eagle RC, Jr. The myofibroblastic component of rubeosis iridis. Ophthalmology. 1983;90(6):721–8.

8. Havens SJ, Gulati V. Neovascular Glaucoma. Dev Ophthalmol. 2016;55:196–204.

9. Pazos M, Traverso CE, Viswanathan A. European Glaucoma Society -Terminology and guidelines for glaucoma, 6th Edition. Br J Ophthalmol. 2025;109(Suppl 1):1–212.

10. Liao N, Li C, Jiang H, Fang A, Zhou S, Wang Q. Neovascular glaucoma: a retrospective review from a tertiary center in China. BMC Ophthalmology. 2016;16(1):14.

11. Lin H, Gao X, Xu Z, Zhang Y, Liao Y, Ren J, et al. The Burden and Clinical Features of Neovascular Glaucoma in a Major Tertiary Care Center in China. J Glaucoma. 2025;34(2):121–6.

12. Lazcano-Gomez G, J RS, Lynch A L NB, Martinez K, Turati M, et al. Neovascular Glaucoma: A Retrospective Review from a Tertiary Eye Care Center in Mexico. J Curr Glaucoma Pract. 2017;11(2):48–51.

13. Olmos LC, Sayed MS, Moraczewski AL, Gedde SJ, Rosenfeld PJ, Shi W, et al. Long-term outcomes of neovascular glaucoma treated with and without intravitreal bevacizumab. Eye. 2016;30(3):463–72.

14. von Elm E, Altman DG, Egger M, Pocock SJ, Gøtzsche PC, Vandenbroucke JP. The Strengthening the Reporting of Observational Studies in Epidemiology (STROBE) statement: guidelines for reporting observational studies. The Lancet. 2007;370(9596):1453–7.

15. Welfare FIfHa. THL -Tautiluokitus ICD-10, Kanta Code Service [cited 2026 29.3]. Available from: https://koodistopalvelu.kanta.fi/codeserver/pages/classification-view-page.xhtml?classificationKey=23.

16. Finland S. Statistics Finland’s free-of-charge statistical databases [Available from: https://pxdata.stat.fi/PxWeb/pxweb/fi/StatFin/StatFinvaerak/statfin_vaerak_pxt_11ra.px/.

17. Holladay JT. Proper method for calculating average visual acuity. J Refract Surg. 1997;13(4):388–91.

18. Virtanen P, Gommers R, Oliphant TE, Haberland M, Reddy T, Cournapeau D, et al. SciPy 1.0: fundamental algorithms for scientific computing in Python. Nature Methods. 2020;17(3):261–72.

19. Davidson-Pilon lsaiPJoOSS, 4(40), 1317, 10.21105/joss.01317.

20. OpenAI. ChatGPT 5 [Large language model] chatgpt.com. 2026.

21. Anthropic. Claude [Large language model] claude.com. 2026.

22. Kelasto. Kela; 2026.

23. Organization WH. World report on vision2019.

24. Rani PK, Sen P, Sahoo NK, Senthil S, Chakurkar R, Anup M, et al. Outcomes of neovascular glaucoma in eyes presenting with moderate to good visual potential. International Ophthalmology. 2021;41(7):2359–68.

25. Kingston EJ, Lusthaus JA. Two-year outcomes of patients presenting to Sydney Eye Hospital with neovascular glaucoma. Int Ophthalmol. 2023;43(8):2763–76.

26. Purola PKM, Ojamo MUI, Gissler M, Uusitalo HMT. Changes in Visual Impairment due to Diabetic Retinopathy During 1980–2019 Based on Nationwide Register Data. Diabetes Care. 2022;45(9):2020–7.

27. Nakano S, Nakamuro T, Yokoyama K, Kiyosaki K, Kubota T. Prognostic Factor Analysis of Intraocular Pressure with Neovascular Glaucoma. J Ophthalmol. 2016;2016:1205895.

28. Strzalkowski P, Strzalkowska A, Göbel W, Loewen NA, Hillenkamp J. Combined vitrectomy, near-confluent panretinal endolaser, bevacizumab and cyclophotocoagulation for neovascular glaucoma -a retrospective interventional case series. F1000Res. 2020;9:1236.

29. Lin H, Gao X, Wu Z, Tam W, Huang W, Dong Y, et al. Treatment modalities and trends for hospitalized patients with neovascular glaucoma: A retrospective study of 10 years. Asia-Pacific Journal of Ophthalmology. 2025;14(1):100136.

30. Inatani M, Higashide T, Matsushita K, Miki A, Ueki M, Iwamoto Y, et al. Intravitreal Aflibercept in Japanese Patients with Neovascular Glaucoma: The VEGA Randomized Clinical Trial. Adv Ther. 2021;38(2):1116–29.

31. Iwasaki K, Kojima S, Wajima R, Matsuda A, Yoshida K, Tsutsui A, et al. Surgical Outcomes of Baerveldt Glaucoma Implant Versus Ahmed Glaucoma Valve in Neovascular Glaucoma: A Retrospective Multicenter Study. Adv Ther. 2025;42(4):1745–59.

32. Kanra AY, Dursun Yilmazşamli T, Altinel MG, İmamoğlu S. Surgical Outcomes of Ahmed Glaucoma Valve in Neovascular Glaucoma Secondary to Diabetic Retinopathy Versus Central Retinal Vein Occlusion. J Glaucoma. 2026;35(3):190–7.

33. StatFin database. Statistics Finland; 2026.

