## Supplementary figures and images for "Neovascular Glaucoma at a Tertiary Centre in Finland, 2008–2024: A Retrospective Cohort Study"

### Supplementary Figure 1

#### Supplementary Figure 1.
#### Annual neovascular glaucoma diagnoses at HUS, 2008–2024

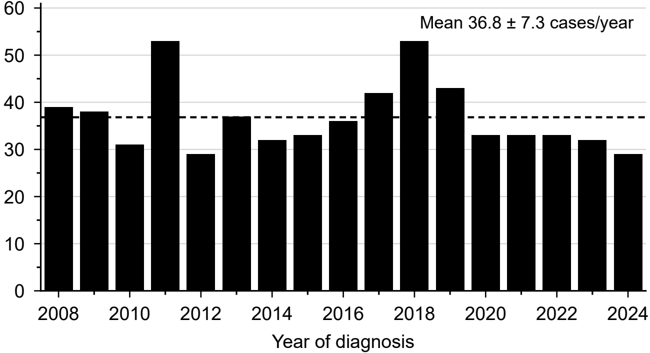
