## Supplementary Figure 2 for "Neovascular Glaucoma at a Tertiary Centre in Finland, 2008–2024: A Retrospective Cohort Study"

**Supplementary Figure 2. Cumulative PRP spots per patient by diagnosis year (3-year centred rolling average, all follow-up)**

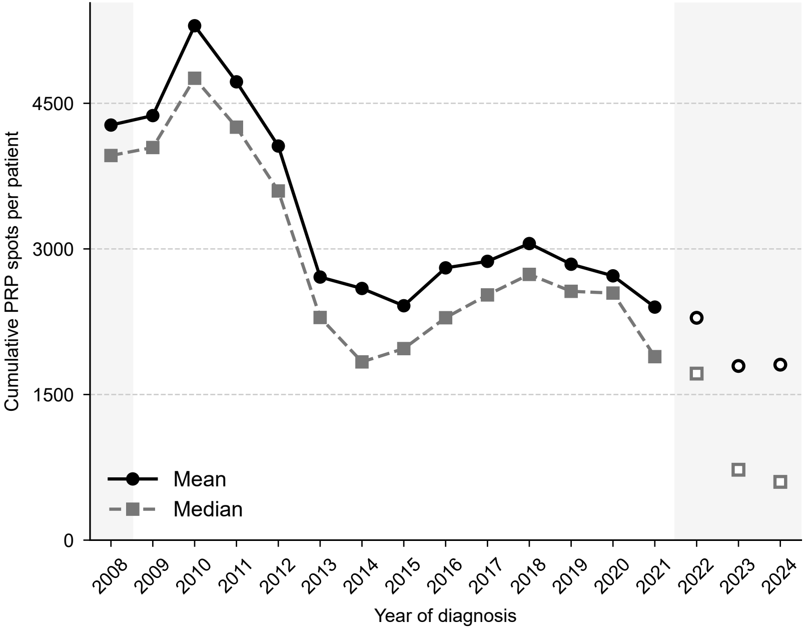
