## Supplementary Methods for "Neovascular Glaucoma at a Tertiary Centre in Finland, 2008–2024: A Retrospective Cohort Study"

Biomicroscopic data included findings on the conjunctiva (hyperaemia); cornea (oedema, Descemet’s membrane folds, neovascularisation); anterior chamber (cells graded according to the SUN scheme (10), depth, and hyphema); anterior chamber angle (most open angle according to Schaffer grading (11), with peripheral anterior synechiae categorised as mild for 0–90°, moderate for 90–270°, and severe for 270–360°; 90° and 270° were classified as moderate); iris (rubeosis or vascular congestion); lens (phakia, cataract, pseudophakia, aphakia, pseudoexfoliation); vitreous (haemorrhage); optic disc (oedema, neovascularisation); and both the macula and peripheral retina separately (haemorrhages, neovascularisation, oedema, and ischaemia). Other diagnosed ophthalmic pathologies were also recorded. The anterior chamber angle value was recorded without indentation. Records of an open angle were classified as gradus 3 and missing synechiae data were categorised as moderate. The biomicroscope data were forward filled to following dates, updating when a new value was recorded.

Visual field data were too sparse after diagnosis for longitudinal analysis. Both Humphrey automated perimetry and Goldmann kinetic perimetry were assessed. For OCT, macular scans acquired with the SPECTRALIS (Heidelberg Engineering GmbH, Heidelberg, Germany) provided sufficient data for analysis; central retinal thickness was extracted from these scans using the Heidelberg Eye Explorer software. Scans with a quality score below 15 or without successful automated segmentation were excluded.
